# Infectious disease modeling for public health practice: projections, scenarios, and uncertainty in three phases of outbreak response

**DOI:** 10.1101/2025.10.20.25338175

**Authors:** Andrew F. Brouwer, Marisa C. Eisenberg, Natalie E. Dean, Harry Hochheiser, Philip Huang, Joseph R. Coyle, Lior Rennert

**Author notes:** **Corresponding author:** Andrew F. Brouwer, Department of Epidemiology, 1415 Washington Heights, University of Michigan, Ann Arbor, MI.

## Abstract

Public health departments need evidence-backed scenario projections to support decision making in infectious disease outbreaks. However, traditional infectious disease models are often not readily deployable or responsive to the urgent questions and priorities of public health departments or health systems. Moreover, uncertainty in model outputs is not always adequately assessed or communicated, potentially undermining trust among public health practitioners and the public. To address these issues, we, the Insight Net Modeling Guidance for Public Health Working Group, used early COVID-19 data from Michigan to illustrate modeling approaches that can be used to answer urgent questions in three key phases of outbreak response: prior to local introduction, early exponential growth, and established transmission with potential interventions. In each phase, we integrate case, hospitalization, and death data and capture ranges of plausible future trajectories. These models, which produce status quo and scenario projections, are intended to inform planning and motivate action rather than forecast precise future outcomes. Importantly, this work offers guidance to focus modeling efforts and provides examples and code for how to fit and implement these models, ultimately serving as both a conceptual guide and practical toolkit to support more transparent, timely, and appropriate use of models in outbreak response.

## Introduction

The first months of the COVID-19 pandemic were full of uncertainty and a need to act in the face of that uncertainty to protect public health and save lives. State, tribal, local, and territorial (STLT) health departments and healthcare systems needed evidence-backed projections and scenarios to support decision making but had little existing in-house expertise and capacity for infectious disease modeling and analytics (let alone the public health resources, workforce, and information technology infrastructure necessary to respond to a pandemic). New government– academic partnerships arose, often in an *ad hoc* manner, to fill this gap.^1^ To improve preparedness for future outbreaks and deepen bonds between health departments and modelers, the Centers for Disease Control and Prevention (CDC) Center for Forecasting and Outbreak Analytics (CFA) was established, along with the National Outbreak Analytics & Disease Modeling Network (Insight Net), a national network of centers comprised of academic, public, and private partners.^2^ As members of the Insight Net Modeling Guidance for Public Health Working Group, our team aims to make high-quality models and analytical tools available for outbreak response.

This article is intended for STLT public health department and healthcare system epidemiologists (referred to as “public health partners”) and their academic or private partners assisting with modeling during an infectious disease outbreak (referred to as “modelers”). In Box 1, we highlight some questions modelers and public health partners can discuss to develop a shared understanding of the goals and expectations of the partnership. In this paper, we illustrate—with accompanying open code—modeling approaches that can be used to urgent questions in each of three key phases of an emerging outbreak: prior to local introduction, early exponential growth, and established transmission with potential interventions. Beyond providing a starting point to avoid the need to reinvent the wheel for each new outbreak, this work aims to help ensure that model outputs are both interpretable and actionable by public health practitioners. Importantly, we emphasize that models do not need to be complicated to be useful: often a relatively simple, back-of-the-envelope analytic approach is sufficient in many cases.

As part of developing shared expectations with public health partners about what questions models are good at answering, it is useful to distinguish between *forecasting*—the prediction of what cases, hospitalizations, and deaths *will be* in the future—and *scenario modeling*—the projection of what these quantities *would be* under the status quo or alternative scenarios. Although the difference may seem subtle, different tools are better suited for each task, and a shared understanding of a model’s goals helps to avoid disappointment or frustration later if, for example, actual cases exceed previous status quo model projections. Because real-world situations may quickly deviate from current conditions, mechanistic infectious disease models may have lower forecasting performance than some statistical or machine learning tools.^3,4^ However, because infectious disease models are mechanistic, they are well-suited to scenario modeling, which can be thought of as *in silico* experiments to answer questions like, “*how bad might outcomes get if we do not intervene?*”, “*are we likely to exceed hospital capacity?*”, and “*how could a given intervention change future trajectories?*” Infectious disease models can be used to design, evaluate, and compare counterfactual scenarios (“what would have happened if…”), as well as retrospectively evaluate the impact of public health interventions. In this article, we focus on scenario modeling, building status quo scenarios based on recent data and illustrating some alternative scenarios.

Our approach also pays careful attention to the inclusion of uncertainty in projections; our goal is to balance the need to bring a number to decision makers with the need to accurately convey how much confidence there is in that number, even if that’s just best-case vs worse-case bounds. It is easy to inadequately assess or communicate uncertainty in the future trajectories projected by infectious disease models, which can reduce both public health partners’ and the public’s confidence in models. By establishing a shared framework, we seek to support more effective, transparent, and responsible use of models in outbreak situational awareness and response.

## Methods

### Overview

Below, we outline three phases of outbreak response that inform our case studies, discuss the data we use to inform our models, describe exponential growth and infectious disease transmission models to project infections, reported cases, hospitalizations, and deaths, and illustrate how we quantify uncertainty in model projections. This paper is geared toward outbreaks with person-to-person transmission and traditional epidemic dynamics, though it may be applicable or adaptable to other pathogens (e.g., vector-borne). These methods may be less applicable to settings where transmission is driven by contact networks of highly clustered susceptibles,^5^ where population immunity is high and introductions often go extinct stochastically,^6^ or where secondary transmission is limited (e.g., point-source, zoonotic). Many technical details are reserved for the Technical Supplement, and all data and code are available in a public repository (see Data Availability). Deidentified, aggregated data for public health surveillance were provided by the Michigan Department of Health and Human Services, and this study was not regulated as human subjects research (University of Michigan Health Sciences and Behavioral Sciences Institutional Review Board HUM00181319).

### Three phases of outbreak response

Data availability and the modeling needs of STLT health departments and healthcare systems change across different phases of outbreak response. Broadly, our goal is to project and quantify uncertainty in reported cases, hospitalizations, and deaths under status quo scenarios. Our models can be extended as needed to include other measures of interest specific to a public health partner’s needs, such as personal protective equipment (PPE), hospital bed capacity, and staffing.

#### Phase 1: Preliminary projections before local spread

Before an expected outbreak has been detected locally, public health partners can use models to understand the potential speed and magnitude of local spread, supporting advanced planning, requests for funding or supplies, and communication with stakeholders. In this phase, exponential growth models adequately capture typical early epidemic growth trajectories. Our models provide preliminary projections based on information from historical or ongoing outbreaks.

#### Phase 2: Local exponential growth

Once local transmission has been sustained for several weeks and the epidemic is growing exponentially, public health partners need local, short-term status quo projections. Our models provide short-term (2-week) exponential projections with parameters informed by recent, local data. In this phase, there is less practical difference between forecasting and scenario modeling, though we still recommend communicating that these projections assume constant epidemic growth rates. We also recommend generating projections for scenarios with increased or decreased epidemic doubling times to illustrate wider uncertainty due to time-varying transmission.

#### Phase 3: Established transmission with potential interventions

If the outbreak continues to spread, public health partners need to consider and evaluate a variety of possible interventions and scenarios to better guide their decision making. Mechanistic infectious disease models are well-suited both to capturing realistic dynamics and uncertainty in status quo projections and projecting alternative scenarios of interest.

### Infectious disease data streams

We rarely have information about the underlying burden of infection. Instead, the data available for modeling is primarily in the form of lists of reported cases—typically those individuals who were symptomatic and sought testing or medical care. These line lists are often simplified to numbers of cases per date. As discussed in the Technical Supplement, we recommend using symptom onset date to capture the infectious disease dynamics. We drop data for recent onset dates (burn-in) because recent data are incomplete due to reporting lags (Figure 1A). (Nowcasting models may be used to reduce or eliminate burn in.^7^) In addition to case data, hospitalizations and deaths are also frequently reported and projected.

**Figure 1.**
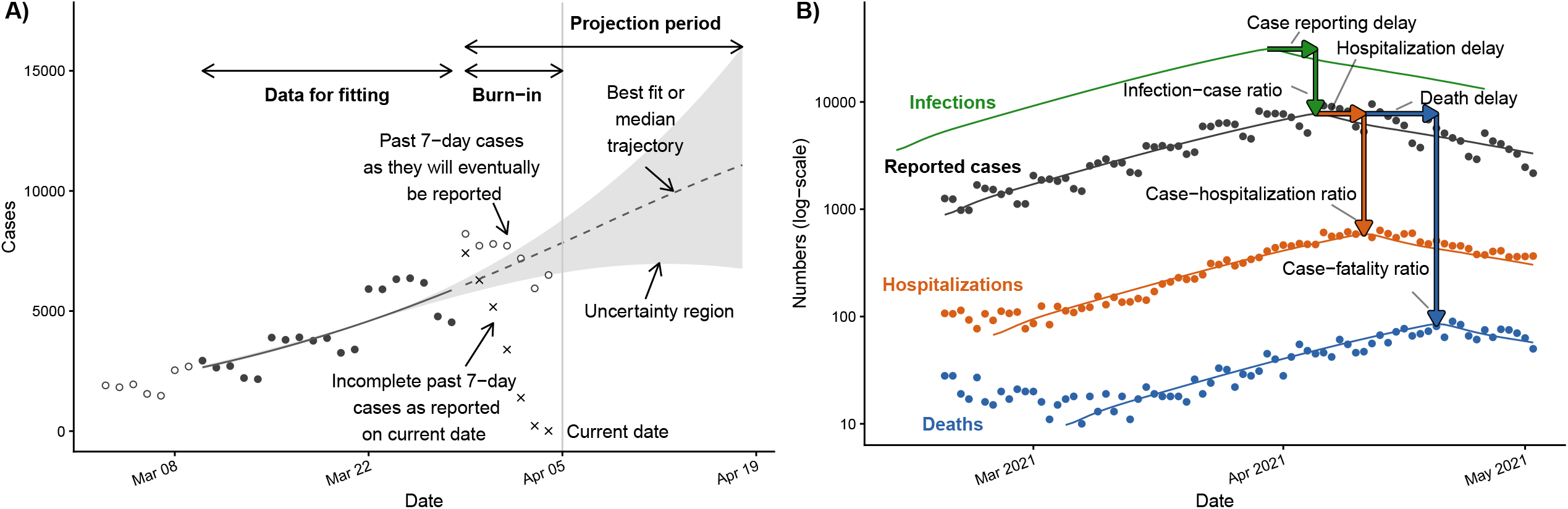
Anatomy of outbreak model projections and supporting data. A) On the current date, case data in the burn-in period (cross points) are incomplete because of reporting delays; they will eventually be filled in (open circles) but are not available on the current date (grey vertical line). The case data within the fitting period (black circles) inform the model projections (dotted line) and uncertainty region (grey ribbon) in the projection period. B) To incorporate data on reported cases, hospitalizations, and deaths and project them and infections, we need to account for both delays and ratios between them. **Figure 1 alt text:** A two-panel figure. The first panel plots cases and a model fit with uncertainty bounds, with multiple annotations highlighting features of the data and model. The second panel plots reported and modeled cases, hospitalizations, and deaths and modeled infections, with annotated arrows highlighting the relationships between the data types.

Our models use six data parameters that describe the relationships between cases, hospitalizations, and deaths: the fraction of infections reported as cases (infection–case ratio) *φ*_*IC*_, the fraction of cases who become hospitalized (case–hospitalization ratio) *φ*_*CH*_, the fraction of cases who die (case–fatality ratio) *φ*_*CD*_, the case reporting lag *τ*_*IC*_, the lag between case reporting and hospitalization *τ*_*CH*_, and the lag between case reporting and death *τ*_*CD*_ (Figure 1B; Table 1). The infection–case ratio cannot be determined from case data alone, so a range of realistic values informed by historic outbreaks is typically considered (it can also be estimated later in the outbreak using population serology). A reasonable range for the case reporting lag can be set from historic outbreaks or from the estimated incubation period and the mean difference between symptom onset and reporting date in the surveillance system. The remaining parameters can be estimated by comparisons of the data streams or, in Phases 2 and 3, through fitting the model to the data. Reporting delays are often exacerbated by high cases numbers that exceed public health workforce capacity.

**Table 1:** Summary of model parameters for COVID-19 in Michigan 2020–21 in each case study across the three outbreak response phases. In Phase 1, we use a best-guess and range for each parameter (with values derived from the literature or expert opinion). In Phases 2 and 3, we fix some data-driven parameters, sample others from a range, and estimate the remaining parameters. Estimated parameters are reported as best fit (Phase 2) or median (Phase 3).

| Parameter | Description | Case Studies |  |  |
| --- | --- | --- | --- | --- |
|  |  | Phase 1 | Phase 2 | Phase 3 |
| *Data parameters (both models)* |  |  |  |  |
| $\varphi_{IC}$ | Infection–case ratio | 0.25 (0.05–0.50) | 0.25 (0.05–0.50) | Est. as $\kappa\varphi_{IC}$: 1.2E6 |
| $\varphi_{CH}$ | Case–hospitalization ratio | 0.10 (0.02–0.20) | Est: 0.08 | Est: 0.075 |
| $\varphi_{CD}$ | Case–fatality ratio | 0.03 (0.01–0.05) | Est: 0.02 | Est: 0.01 |
| $\tau_{IC}$ | Case reporting delay (days) | 6 (4–10) | 6 (4–10) | 6 (4–10) |
| $\tau_{CH}$ | Hospitalization delay (days) | 6 (4–10) | 6 | 6 |
| $\tau_{CD}$ | Death delay (days) | 15 (10–20) | 15 | 15 |
| *Exponential growth model parameters* |  |  |  |  |
| $C(0)$ | Initial number of cases | $\varphi_{IC} \times 2^{\tau_{IC}/\delta}$ | Est: 1.3E3 | — |
| $\delta$ | Doubling time (days) | 3.5 (3.0–4.0) | Est: 17.2 | — |
| *Infectious disease transmission model parameters* |  |  |  |  |
| $R$ | Reproduction number | — | — | Est: 1.71 |
| $\sigma$ | Latency rate (1/day) | — | — | (1/8, 1/4) |
| $\gamma$ | Recovery rate (1/day) | — | — | (1/10, 1/5) |
| $I_0$ | Infectious fraction at time 0 | — | — | (1E-6, 1E-1) |
| $\kappa$ | Size of the at-risk population | — | — | Est. as $\kappa\varphi_{IC}$: 1.2E6 |

### Exponential growth model

In Phases 1 and 2, when case numbers approximately follow exponential growth, we use an *exponential growth model*. This class of model may be thought of as informed back-of-the-envelope calculations for generating short-term, status quo scenario projections, as real, longer-term dynamics will deviate from these models.

We model numbers of actual infections *Y*(*t*), reported cases *C*(*t*), hospitalizations *H*(*t*), and deaths *D*(*t*) as functions of time since outbreak start *t*. In addition to the data parameters described above, these models are informed by the initial number of cases *C*(0) and the epidemic doubling time *δ* (Table 1).

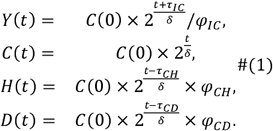

The model in Eqs (1), which captures exponential growth in cases and the delays and ratios between the data streams, is used to project *Y*(*t*), *C*(*t*), *H*(*t*), and *D*(*t*) for each day *t*.

### Infectious disease transmission model

In Phase 3, we use a compartmental *infectious disease transmission model*, based on a susceptible–latent–infectious–recovered (SLIR) framework and simulated with differential equations.^8–10^ The model describes the fraction of the population in each compartment (Figure 2), accounting for the effective transmission rate (*β*), the rate of transition from the latent to infectious phase (*σ*), and the recovery rate from the infectious compartment (*γ*). The effective reproduction number *R*—the number of secondary cases generated by a single infectious individual in the current population—here equal to *β*/ *γ*, is more intuitive than the transmission rate: we recommend reporting the effective reproduction number rather than *β* when communicating with public health partners or the public, and, to that end, we report *R* rather than *β* in Table 1. The system of differential equations for the model is given in Eqs (2). As discussed in more detail in the Technical Supplement, we include more realistic durations for the latent and infectious states by modeling each as two consecutive compartments, each with half the total duration (and hence twice the rate in Eqs (2)).^11^

**Figure 2.**
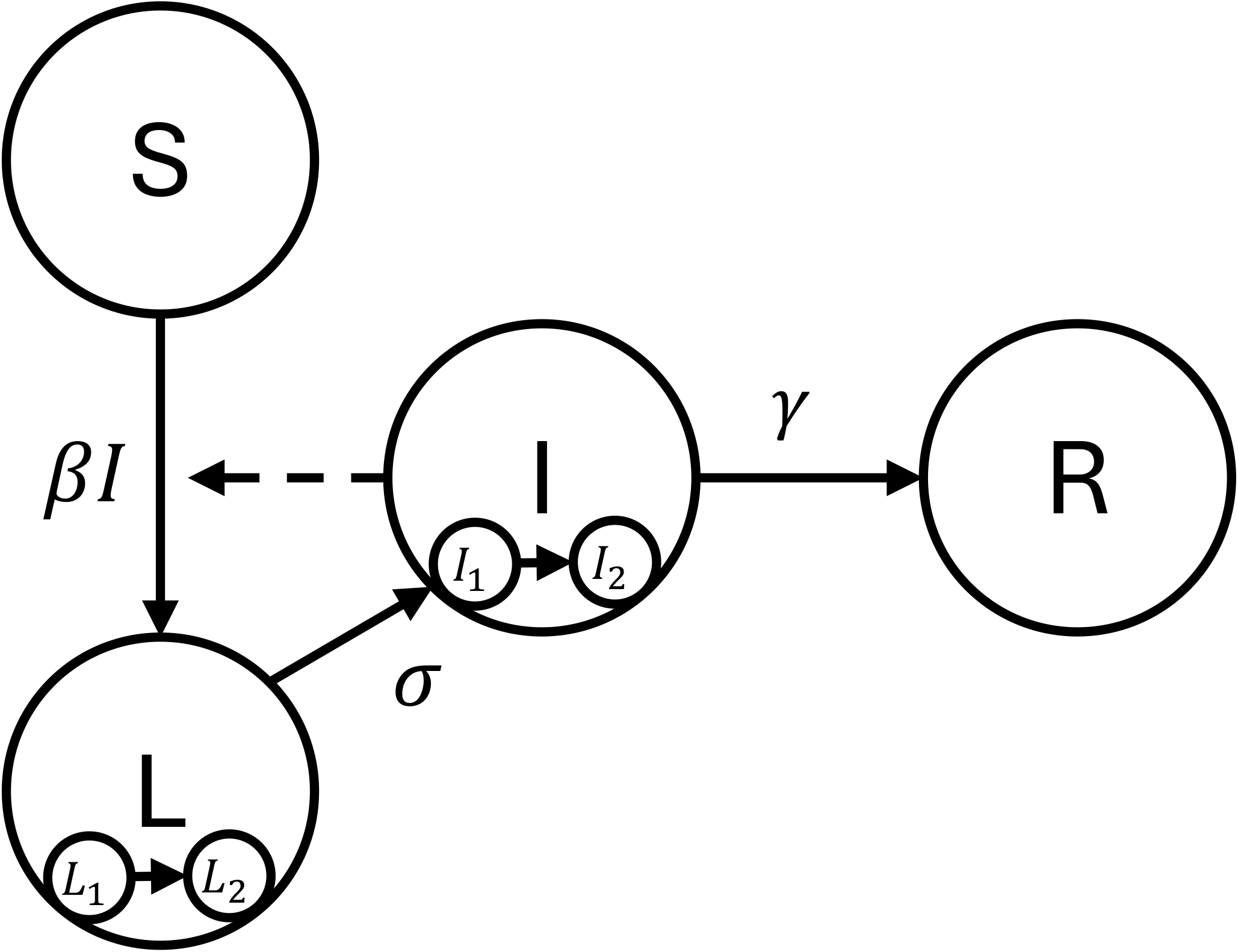
A compartmental infectious disease model with susceptible (S), latent (L), infectious (I), and recovered (R) compartments. The model accounts for the transmission rate (β), the latency-to-infectiousness rate (σ), and the recovery rate (γ). To create realistic distributions of latent and infectious periods, the latent and infectious phases are each modeled as two consecutive compartments (distributed delay). This model can be expanded to account for mechanisms or data streams specific to the disease of interest. **Figure 2 alt text:** A schematic of a compartmental infectious disease model, with arrows describing flows between compartments.

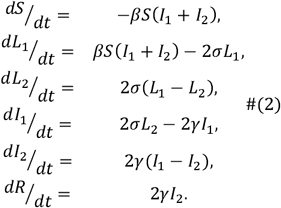

The model initial conditions are described in the Technical Supplement, as a function of the fraction of individuals that are initially infectious, *I*_0_.

To connect the model to data and generate projections for numbers of actual infections *Y*(*t*), reported cases *C*(*t*), hospitalizations *H*(*t*), and deaths *D*(*t*) over time *t*, we again account for the data delay and ratio parameters. Additionally, we need the size of the at-risk population *κ* (not the catchment population, which may include a substantial recovered fraction or otherwise not be at risk of infection in the short-term) to translate the simulated *fraction* infectious into *numbers* of infectious individuals. Importantly, the values of the size of the at-risk population *κ* and the infection–case ratio *φ*_*IC*_ cannot be separately determined from case data. The observed case trajectory could be explained equally well by a larger at-risk population paired with a smaller infection–case ratio or by a smaller at-risk population paired with a larger infection–case ratio. There would be more unreported cases in the first situation, but without other data, e.g., serosurveys, we cannot determine which situation is correct. Accordingly, *κφ*_*IC*_ is treated as a single, estimated parameter. For brevity, the connection between the Eqs (2) and *Y*(*t*), *C*(*t*), *H*(*t*), and *D*(*t*) is given in the Technical Supplement.

### Fitting models to data

In Phases 2 and 3, we fit models to local data using parameter estimation. We assume that daily reported cases, hospitalizations, and deaths are drawn from Poisson distributions with means *C*(*t*), *H*(*t*), and *D*(*t*), respectively. The negative log likelihood *NLL*_*C*_(*θ*) of observing the reported cases *z*_*C*_(*t*) on each day *t* in the set of days included for fitting cases {t_*C*_} is given in Eq (3). In lay terms, this equation measures the discrepancy between the modeled cases *C*(*t*) and the observed cases *z*_*C*_(*t*), so that we can minimize that discrepancy by adjusting the parameters *θ*.

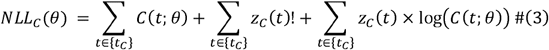

The ! in Eq (3) denotes the factorial function. The negative log-likelihoods for hospitalizations and deaths are defined analogously, using hospitalization and death data, *z*_*H*_(*t*) and *z*_*D*_(*t*), and their data periods {*t*_*H*_} and {*t*_*D*_}, respectively. Further details are given in the Technical Supplement. The total negative log-likelihood is the sum of the three negative log-likelihoods: *NLL*(*θ*) = *NLL*_*C*_(*θ*) + *NLL*_*H*_(*θ*) + *NLL*_*D*_(*θ*). We find the best fit parameters *θ* by minimizing the negative log-likelihood *NLL*_*C*_(*θ*). In our case studies, below, we use the `optim` function in R v4.4 with the Nelder-Mead algorithm, a classic and widely used parameter estimation approach.

### The importance of quantifying and communicating uncertainty

Although decision makers often like the simplicity of a single number (e.g., the best-fit prediction), we recommend providing an estimate of the degree of uncertainty in that number. Indeed, inappropriately confident predictions can reduce trust in models and modelers, as was highlighted in the sensational overestimates given in the 2014–15 Ebola outbreak^12,13^ and more recently in the COVID-19 pandemic.^14^ Even relatively crude approaches to uncertainty, giving best and worst case scenarios can be extremely helpful in providing intuition to decision makers. We want to be able to ask questions like, “*What range of dynamics is possible between the best and worst cases?*” and, “*Is substantial action needed to avoid even the best-case scenario?*”

There are several sources of uncertainty in outbreak analytics and modeling. One source is noise in the data itself. Another source comes from not knowing the current underlying state of the system (e.g., the actual number of infected individuals) or the values of model parameters (e.g., the reproduction number or case reporting fraction). A state in the model is *observable* if it can be determined from the available data. In an infectious disease outbreak, the numbers of reported cases, hospitalizations, and deaths are observable, but the number of infections is typically not. A model parameter is *identifiable* from the data if there is a unique, best-fit value of that parameter associated with the data. For example, the case–fatality ratio is typically identifiable (as we observe both cases and deaths), but the infection–case ratio and the case reporting lag will not be (as we do not observe infections). There is a rich literature on identifiability and observability of parametric models,^15–18^ and we are only scratching the surface here.

In an outbreak—and particularly during the growth phase of an outbreak—many relevant parameters will not be identifiable from the available data. However, projections may depend on the specific values of unidentifiable parameters values or unobservable states, and so we may have to rely on external information, such as estimates from other outbreaks, similar pathogens, or even just reasonable ranges determined by the modeler or other expert opinion.

There are many approaches to uncertainty analysis, and we will use a different approach in each of the three Phases of the outbreak. In Phase 1, there is no local data, and our uncertainty will be based only on reasonable ranges of the parameters. Using Sobol’ sampling,^19,20^ we sample a large number of combinations of parameters from across their ranges, generate projections for each combination, and calculate credible intervals for each day in the projection. (Please see the Technical Supplement for a discussion of the number of samples needed). In Phase 2, we use reasonable ranges for the parameters that are not identifiable, and we will create reasonable ranges for the identifiable parameters consistent with the noise in the data. For this case study, we will use +/-25% of the best-fit parameter values, which produces confidence intervals capturing the level of noise in our data, but a larger range may be appropriate for capturing noisier data. As in Phase 1, we sample a large number of combinations of parameters from across their ranges, generate projections for each combination, and calculate quantiles for each day in the projection

The mechanistic model in Phase 3 has a larger number of unidentifiable parameters that cannot be uniquely estimated from the data but can affect the projection trajectory. To overcome this difficulty, we will use a two-step combination of sampling and estimation. The general idea is that we generate a large number of combinations of reasonable values for the unidentifiable parameters in the first step (Figure 3A), and then for each of these combinations, we estimate the identifiable parameters using maximum likelihood estimation as discussed in the previous section (Figure 3B).^21^ To ensure that each of these parameter sets reasonably fit the data, we drop parameter sets whose NLLs exceed a threshold above the minimum NLL. (Please see the Technical Supplement for discussion of this threshold). We then generate a projection for each of the remaining parameter combinations (Figure 3C) and plot the bounds for the 50% and 95% credible intervals (Figure 3D), indicating a region of more likely projections and the range of best- and worst-case projections. (Figure 3C and D also represent how uncertainty is estimated for Phases 1 and 2).

**Figure 3:**
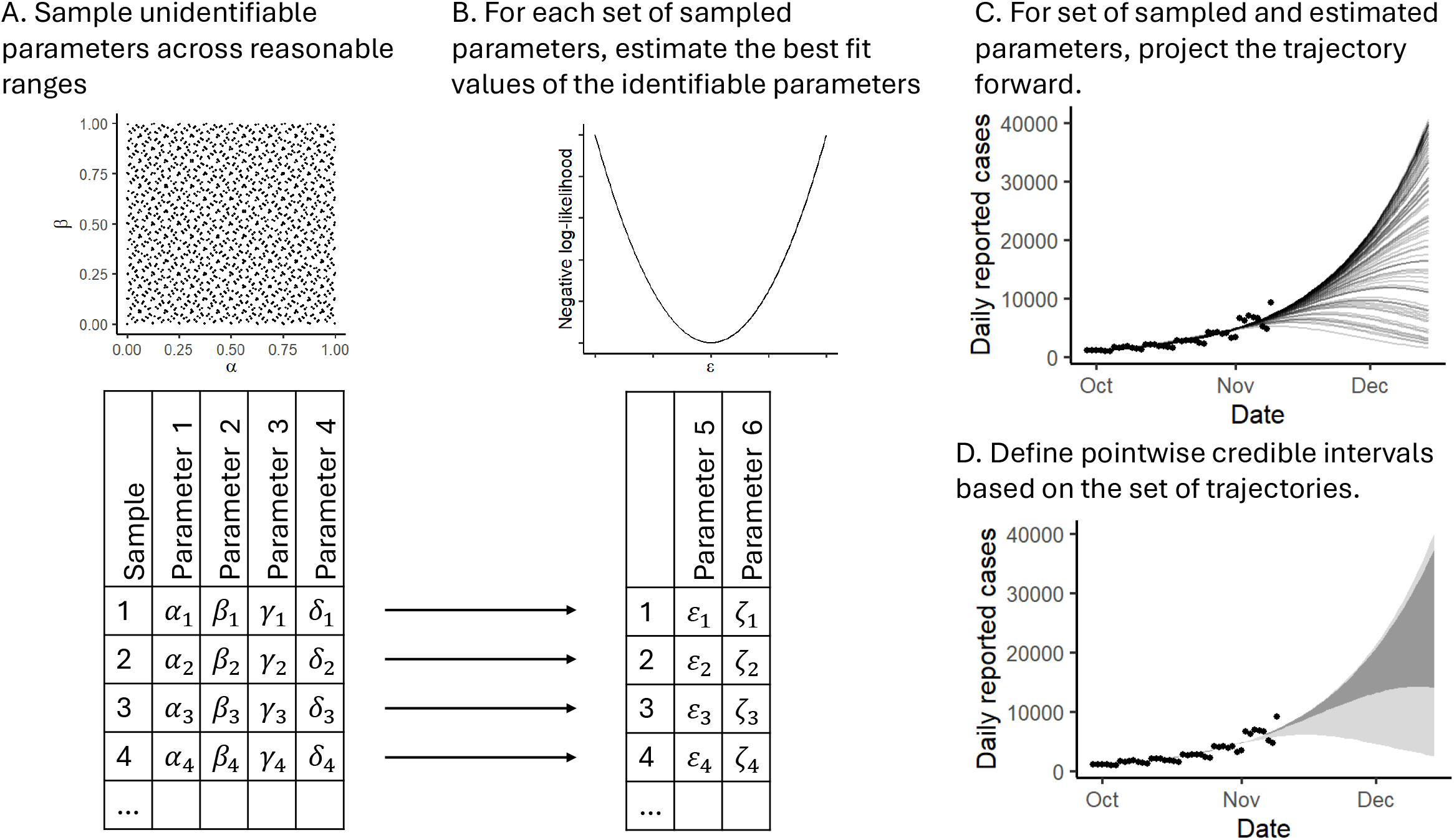
Handling uncertainty for mechanistic infectious disease model forecasting. We use a hybrid sampling–estimation approach to sample unidentifiable model parameters and estimate identifiable model parameters. A) We generate many combinations of reasonable values for the unidentifiable parameters. B) For each combination of unidentifiable parameters, we estimate the values of the identifiable parameters by maximizing the fit (minimizing the negative log-likelihood). C) We generate a projection for each combination of sampled and estimated parameters that reasonably fits the recent data. D) We display 50% and 95% credible intervals for the projected trajectories. This approach provides a full array of possible future trajectories consistent with the recent past. **Figure 3 alt text:** A figure with four panels. A) A graph of sampled values of two parameters demonstrating coverage over the parameter space and a table of sets of sampled parameters. B) A graph of a negative log-likelihood indicating a maximum-likelihood best fit and a table of sets of estimated parameters. The tables of sampled and estimated parameters are connected by arrows indicating row-wise connections. C) A graph of case data and a large number of infectious disease trajectories that all fit the case data but have different projections. D) A graph of case data and two uncertainty regions.

### Case studies

We illustrate our modeling approaches in each of the three Phases using COVID-19 case, hospitalization (new confirmed and probable COVID-19 admissions), and death data from Michigan, 2020–21.

#### Phase 1: Preliminary projections before local spread

##### Approach

The modeling goal of Phase 1 is to understand the potential speed and magnitude of local spread, supporting advanced planning and communication with stakeholders. To illustrate the Phase 1 approach, we use the best-guess parameter values and ranges in Table 1, starting from a single infected individual, and provide six-week projections.

##### Results

On the day six weeks after introduction, we project 4,100 (50% CI: 2,400–7,800; 95%CI: 1,500–15,100) reported cases; 54,000 (50% CI: 33,000–154,000; 95%CI: 12,000– 653,000) actual infections; 120 (50% CI: 60–190; 95%CI: 20–550) hospitalizations, and 6 (50% CI: 3–11; 95%CI: 1–33) deaths (Figure 4).

**Figure 4:**
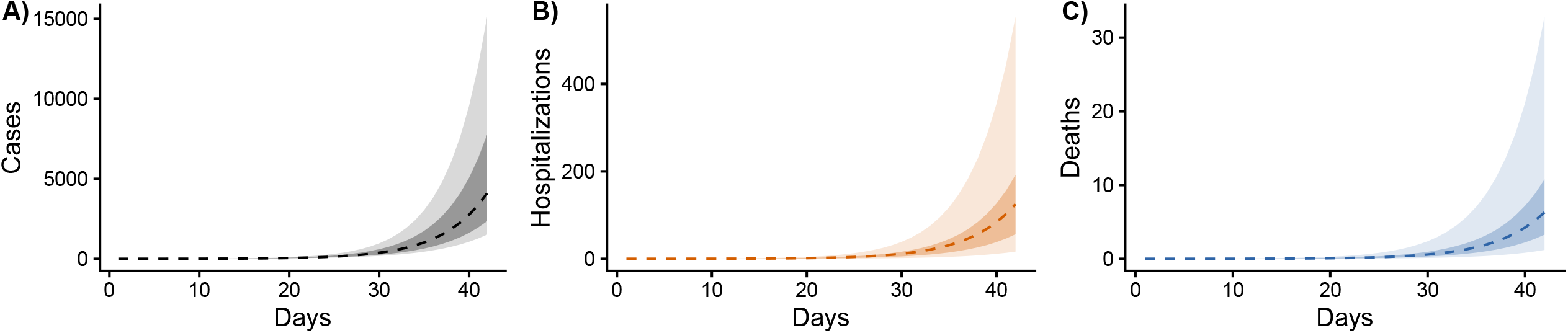
Phase 1 (before local transmission) projections of A) reported cases, B) hospitalizations, and C) deaths. The x-axis is days after a hypothetical first infection. The dotted lines represent best-guess projections, the lighter ribbons denote 95% credible intervals, and the darker ribbons denote 50% credible intervals. **Figure 4 alt text:** A figure with three panels, each with a projected trajectory and ribbons denoting uncertainty regions.

#### Phase 2: Local exponential growth

##### Approach

The modeling goal of Phase 2, once local circulation has begun, is to provide short-term projections to guide planning. To illustrate the Phase 2 approach, we fit our exponential models to the four weeks of data preceding November 1, 2020 (chosen as a typical example of epidemic growth when all data streams were available), using a 1-week burn-in for the case data, and make a two-week status-quo projection. In this phase, projections beyond 2-weeks are generally unreliable because of how quickly real dynamics can change and deviate from exponential models. As summarized in Table 1, we fix *τ*_*CH*_ and *τ*_*CD*_ to data-driven values and only estimate the initial case counts, doubling time, case–hospitalization ratio, and case–fatality ratio (*C*_0_,*t, φ*_*CH*_, and *φ*_*CD*_). Note that the infection–case ratio and delay (*φ*_*IC*_, *τ*_*IC*_) are not identifiable from the available data, so we do a sensitivity analysis, using the best-guess estimates and ranges from Phase 1 to estimate infections (included in the code but not shown in the results).

##### Results

On the day two weeks after November 1, 2020, we project 6,600 (50%CI: 5,400–8,300; 95%CI: 4,100–12,000) reported cases, 400 (50%CI: 330–500; 95%CI: 230–730) hospital

admissions, and 60 (50%CI: 50–73; 95%CI: 36–100) deaths (Figure 5). The best-fit doubling time in this period was 17.2 days, much slower than the early initial spread in Phase 1.

**Figure 5:**
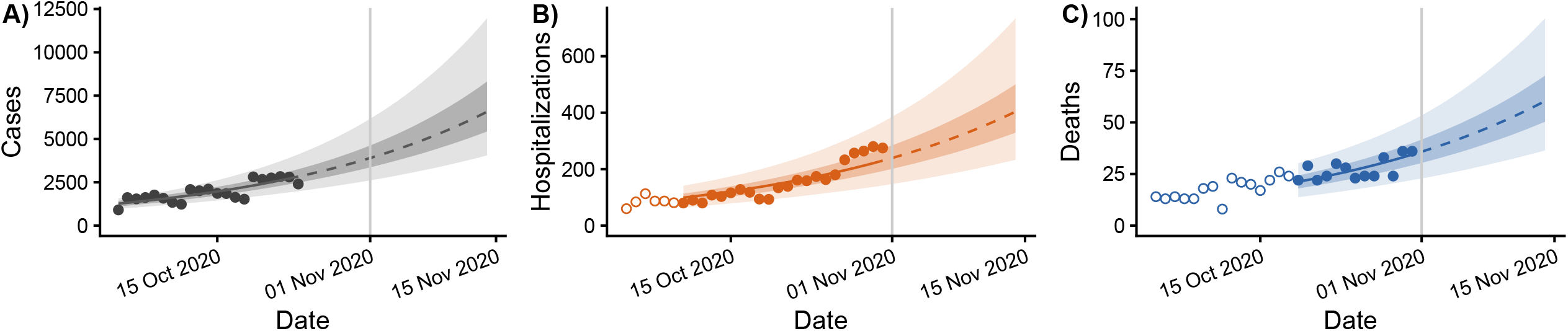
Phase 2 (local exponential growth) projections of A) reported cases, B) hospitalizations, and C) deaths. The solid and dotted lines represent median projections in the data and projection periods, respectively, the lighter ribbons denote 95% confidence intervals, and the darker ribbons denote 50% confidence intervals. Case data are burned for 1 week prior to current date (November 1, 2020; vertical grey line) to account for reporting delays from symptom onset. Hospitalization and death data are delayed relative to cases, so we do not include data that do not correspond to the case data that inform the model fit (open points). **Figure 5 alt text:** A figure with three panels, each with data and a projected trajectory and ribbons denoting uncertainty regions.

#### Phase 3: Established transmission with potential interventions

##### Approach

The modeling goal of Phase 3 is to make projections accounting for dynamics other than exponential growth and to evaluate potential interventions and scenarios. To illustrate the Phase 3 approach, we fit our models to the six weeks of data preceding April 5, 2021 (chosen as a nice example of how scenario models can be revisited in hindsight) and make four-week status-quo projections. As summarized in Table 1, parameters *τ*_*CH*_ and *τ*_*CD*_ are fixed as in Phase 2. The latency (*σ*) and recovery rate (*γ*) parameters and case reporting delay *τ*_*IC*_ are sampled from ranges. The initial fraction infectious, *I*_0_, is not observable, and we sample it from a large range of possible values (10^−6^, 10^−1^). For given set of fixed and sampled parameters, we estimate identifiable parameter combinations *R, κφ*_*IC*_, *φ*_*CH*_, and *φ*_*CD*_.

We illustrate alternative scenario modeling first by simulating a ±25% change to the reproduction number starting on April 5. This range is intended to capture realistic changes or trends in dynamics not tied to specific events (e.g., caused by changing contact patterns); the width of this range can be decided by consensus between modelers and public health partners. We recommend routinely preparing both status quo scenarios and scenarios with greater or lower transmission to demonstrate the additional uncertainty in projections caused by time-varying parameters.

Scenarios can also be used to estimate the potential impact of larger changes, such as stay-home orders or social distancing fatigue. Scenario projections can also be revisited weeks later to see how consistent actual dynamics were with each of the scenarios considered. Here, we compare the data as eventually reported to a scenario with a 45% reduction in reproduction number, which closely matches the data.

##### Results

We estimated the reproduction number in this period to be 1.71 (95% CI: 1.50, 2.01). On the day four weeks after April 5, 2021, we project a median 12,000 (50%CI: 7,400–19,000; 95%CI: 4,200–30,400) reported cases, 890 (95%CI: 620–1,250; 95%CI: 400–1,740) hospital admissions, and 116 (50%CI: 93–141; 95%CI: 69–168) deaths (Figure 6A–C). A wide range of future trajectories—from continued explosive growth to susceptible burnout and epidemic decay—are plausible given the recent epidemic growth trajectory, highlighting the difficulty of epidemic forecasting given the data that are typically available.

**Figure 6:**
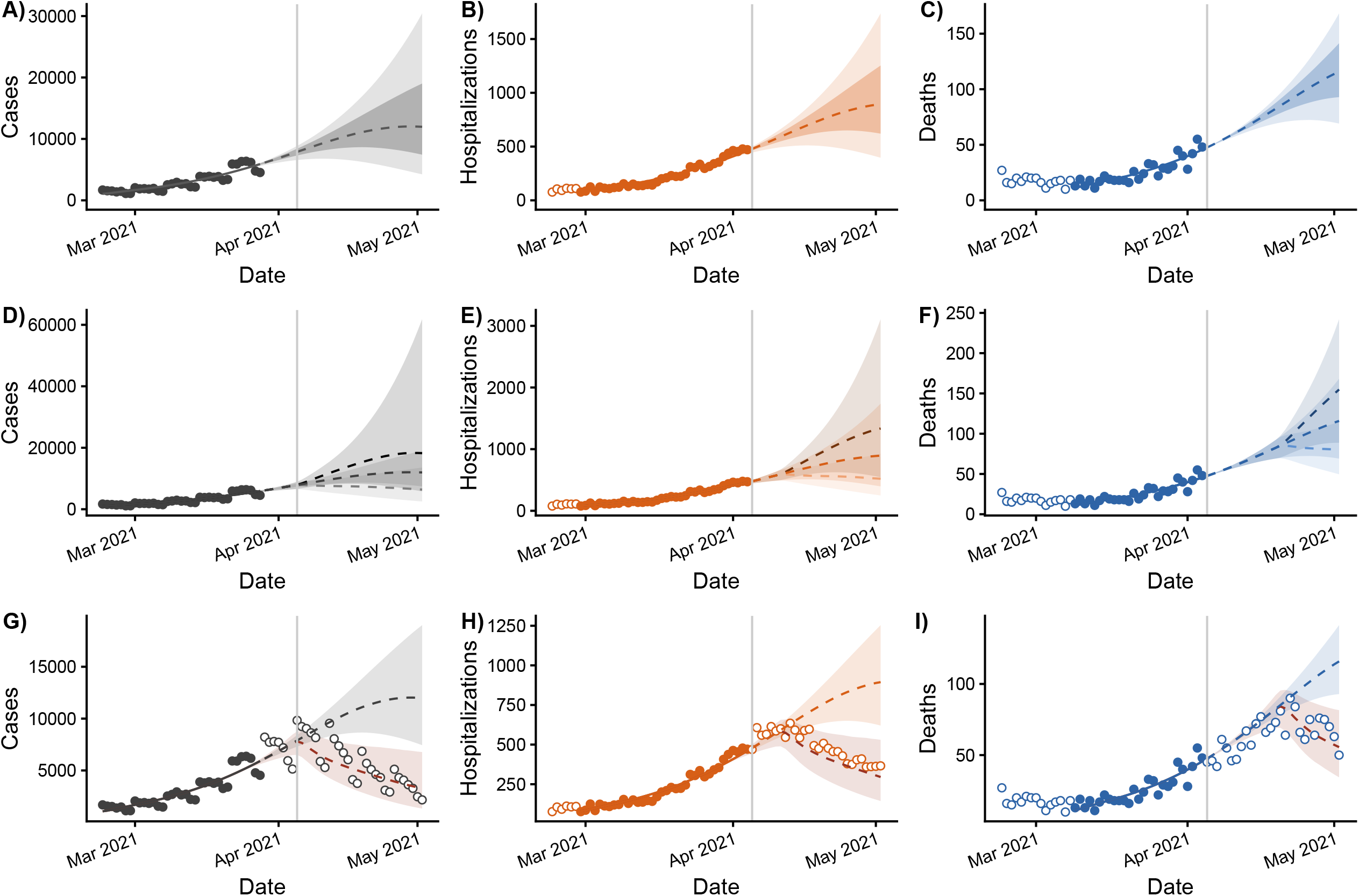
Phase 3 (interventions and scenarios) projections of reported cases, hospitalizations, and deaths. Status-quo projections are given in subfigures A–C, ±25% reproduction number scenarios in D–F, and 45% decreased reproduction number scenario, retrospectively compared to the eventually reported data in G–I. The lines represent median projections, the lighter ribbons denote 95% confidence intervals, and the darker ribbons denote 50% confidence intervals. Case data are burned for 1 week prior to current date (vertical grey line) to account for reporting delays. Hospitalization and death data are delayed relative to cases, so we do not include data (open points) that do not correspond to the case data that inform the model fit. **Figure 6 alt text:** A figure with nine panels, each with data and a projected trajectory and ribbons denoting uncertainty regions.

Under the scenario with a 25% increase in the reproduction number (median 2.14), we project a median 18,200 reported cases, 1,330 daily hospital admissions, and 155 deaths, and, under the scenario with a 25% decrease in the reproduction number (median 1.28), we estimate a median 6,400 reported cases, 520 daily hospital admissions, and 80 deaths (Figure 6D–F). In retrospect, the scenario with a 45% decrease in the reproduction number (median 0.94) is consistent with the cases, hospitalizations, and deaths as they were eventually reported (Figure 6G–I).

## Discussion

We developed models to project infections, cases, hospitalizations, and deaths, along with uncertainty estimates, under status quo assumptions in three phases of outbreak response: before local transmission, local exponential growth, and established transmission with potential interventions. These projections are not forecasts and are not likely to come to pass as is. Instead, they provide information about the potential magnitude of the problem if nothing changes. And change is likely: either through action coordinated by STLT health departments or simply as a function of behavior change by the public in response to news. Our projections provide a benchmark for decisions makers to determine if and what action to take, such as planning for hospital capacity, applying for aid for additional protective equipment, starting early surveillance to catch silent circulation prior to hospitalizations or deaths, and building support for larger interventions (e.g., health campaigns, vaccination clinics, or stay-at-home orders).

As we see here, even relatively simple analytic approaches (exponential models) can be useful for public health practitioners if thoughtfully applied. Although primarily inspired by work on pandemic and seasonal respiratory viruses, our models are intended to be generic and flexible enough to fit many outbreaks, as long as the dynamics can be reasonably approximated by exponential growth or SLIR models. These models are starting points that can (and will likely need to) be modified for specific outbreaks, and they will not be appropriate for all types of outbreaks, as discussed earlier. Additionally, these models are not intended as a theoretical exercise; they need to be responsive to the specific needs of STLT public health departments and healthcare systems. To be responsive, modelers may need to add projections of other quantities of interest; for the COVID pandemic, these projections could have included intensive care unit beds, PPE needs, or extracorporeal membrane oxygenation (ECMO) machines. Public health partners may also be interested in projections for finer geographic subdivisions (e.g., county), age group, race and ethnicity group, or other subgroups of interest. Ultimately, modelers need to be in close conversation with public health partners (Box 1), not only to deliver responsive modeling but also to effectively and accurately communicate those estimates and their uncertainties.

We again emphasize that these model projections should not be thought of as forecasts, meaning that they are not projecting cases, hospitalization, and deaths as we expect them to be. Deviation from the projections does *not* mean that the models were wrong. Instead, reality may be deviating from the status quo assumptions of the model, particularly through time-varying parameters (e.g., changing transmission rate or case–fatality ratio, etc). Rather than treating the status quo model projections as forecasts, modelers can provide projections from a range of possible scenarios (Figure 6D–F). It is then possible to see, in retrospect, which scenario was most consistent with what occurred (Figure 6G–I). One challenging aspect of providing these modeling projections that we did not discuss in detail is how to make serial projections while underlying parameters change, e.g., as interventions are implemented or as the public’s behavior changes. One could incorporate changing dynamics into the model directly, possibly informed by previous scenario modeling. For example, in Figure 6G–I, we might take the 45% reduction in the reproduction number scenario as the new status quo from which to make subsequent status-quo and scenario projections. Alternatively, parameters can themselves be modeled as time-varying, as step functions, splines, or using other functional forms.

Our approach to uncertainty encourages modelers and public health partners to avoid overconfidence in best-fit projections because the range of trajectories that reasonably fit recent data may have a wide spread of future dynamics. There are multiple combinations of unidentifiable parameters, the population at risk, and fraction of the population that is infectious that are consistent with recently observed data (Figure 6A–C). It is also important to communicate with public health partners or to the public that the uncertainty bounds are for status quo scenarios and do not include the possibility of changes in transmission or other parameters and that they do not represent forecasts; pairing the status quo scenario projections with alternative scenario projections can help to make this distinction clear.

In our collective experience as part of Insight Net and as academic modelers or public health professionals, we recognize that STLT public health departments and healthcare systems rarely need complex modeling techniques, especially in the early days of an outbreak. Instead, educated guesses and “back-of-the-envelop” calculations can be more effective when they are fast and responsive and easy to communicate. Here we have provided modeling approaches, with associated open code, to serve as what we hope to be an accessible starting point for a wide variety of both modelers and public health partners.

## Supporting information

Technical Supplement

## Acknowledgments

We thank Juliet Pulliam for her contributions to the early discussions of this work, which helped shape the case studies. We thank the members of the Insight Net Modeling Guidance for Public Health Working Group for providing critical feedback. We thank Annie Niu, Troy Zhou, and Stephanie Iorga for reviewing and testing the code.

## Data Availability

All data and code have been made available in a Zenodo repository at https://doi.org/10.5281/zenodo.17253531.

## Box

**Box 1:** Questions to build shared understanding between modelers and public health partners.

- What are the key questions to answer at this time, and how will we decide when different questions become more important?
- What are the key metrics that we want our models to project?
- What is the difference between forecasting and scenario modeling?
- What data are available for the model, and what are the strengths and limitations? Is a data sharing agreement needed?
- What assumptions does the model make?
- Why might the outbreak deviate from the model projections?
- What alternative scenarios do we want to consider?
- Why is it important to estimate uncertainty, and what degree of certainty is useful?
- What does a “reasonable” parameter range or “reasonable” fit to the data look like?

