## Supplementary material for "Infectious disease modeling for public health practice: projections, scenarios, and uncertainty in three phases of outbreak response": Technical Supplement

### Modeling considerations for dates in infectious disease data

There are multiple dates that may be associated with each case in an infectious disease surveillance system: *reporting date* is the date that the case is reported to the system, *testing date* is the date that a sample was collected from the individual, and *onset date* is the date that individual developed symptoms. Each of these dates have pros and cons from the perspective of plotting an epidemic curve, i.e., the curve of cases per date. When epidemic curves are drawn by ***reporting date***, we simply plot the number of cases reported on that day. However, this approach is highly vulnerable to administrative errors, such as bulk reporting of case backlogs (from misplaced samples in labs, correctional facilities, etc.) or temporary underreporting (from reporting systems being unavailable, etc.), and these errors can substantially impact model fitting and projections. Additionally, because there can be large variations in the time between symptom onset and testing and between testing and reporting, reporting date can misrepresent the true dynamics of epidemic curve. Our approach can be modified to work with reporting date, but serious thought should go into how unusual reporting dates are handled.

Instead, drawing the epidemic curve by ***date of symptom onset*** is more likely to capture the underlying infectious disease dynamics accurately. However, using onset date means that previous days' data must be revised each day with newly reported cases being assigned to earlier onset dates. Thus, from the point of view of the current date, a true increase in cases may look like a plateau, or a plateau may look like a decrease, simply because individuals with recent infections haven't yet had time to develop symptoms, test, and get reported to the surveillance system (Figure 1a). Accordingly, for model fitting and projections, it is important to not include onset data from the most recent dates; in practice, that typically looks like not including the past 7 days of day, although this number can change depending on the pathogen and reporting system. As an advanced option, you may consider reducing or eliminating this burn-in period by using statistical or machine learning "nowcasting" models. These models extrapolate data trends to fill in the expected number of cases not yet reported due to lags.

In addition to lagged reporting, there may be additional artifacts of data collection (e.g., the case data, such as in Figure 4, display a strong day-of-the-week pattern). It is important to understand the source of these artifacts and decide when the data can be used as is (e.g., the day-of-the-week pattern gets averaged out over multiple weeks) or when they ought to be adjusted for (e.g., reporting delays).

### Additional details for the infectious disease transmission model

Our model is an extension of the SIR model that accounts for a latent infection period in which an individual is infected but not yet infectious. Note that the latent compartment has also been called an "exposed" compartment in the literature, but this terminology has more recently fallen out of favor since "latent" is a more accurate description of an infected-not-yet-infectious person than "exposed". One limitation of the basic SIR model is the implicit assumption of an exponential infection length, where most infectious periods are very short and a few are very long (mode $\ll$ mean). In reality, the distributions of the latent and infectious periods typically clustered around the average value (mode $\approx$ mean). For example, the majority of people infected with SARS-CoV-2 are infectious for approximately 8–12 days. To create a more realistic distribution of the latent and infectious periods, we model the latent and infectious periods are each modeled as two consecutive compartments, each with half the expected duration (i.e., twice the transition rate). This approach is known as a distributed delay or linear chain trick; in brief, replacing one compartment with exit rate $\alpha$ (and mean duration $1/\alpha$) with $n$ compartments each with exit rate $n\alpha$ (and thus the same total mean duration of $n/n\alpha = 1/\alpha$) changes the distribution of residence times from exponential with rate parameter $\alpha$ to an Erlang (or, more generally, gamma) distribution with rate parameter $n\alpha$ and shape parameter $n$. Finally, because we are focused on short-term projections, we do not consider waning immunity (i.e., a transition form Recovered to Susceptible).

The system of differential equations for the mechanistic infectious disease transmission model is given in Eqs (S1) (Eqs (2) in the main text).

$$
\begin{aligned}
{dS}/{dt} &= -\beta S(I_1 + I_2),\\
{dL_1}/{dt} &= \beta S(I_1 + I_2) - 2\sigma L_1,\\
{dL_2}/{dt} &= 2\sigma(L_1 - L_2),\\
{dI_1}/{dt} &= 2\sigma L_2 - 2\gamma I_1,\\
{dI_2}/{dt} &= 2\gamma(I_1 - I_2),\\
{dR}/{dt} &= 2\gamma I_2.
\end{aligned}
\tag{S1}
$$

For any given simulation, we only model the at-risk population the simulation start date, so that the initial number of recovered $R(0)$ is 0. This choice alleviates the need to estimate the longer-term dynamics of the system, but it does create additional uncertainties for the initial conditions of the latent and infectious compartments. In practice, we treat the number of infectious people at the start time, $I_0$, as a parameter and set the initial conditions of the compartments using quasi-steady state approximations. Specifically, we set

$$
\begin{aligned}
I_1(0) &= I_0/2,\\
I_2(0) &= I_0/2,\\
L_2(0) &= I_1(0)\gamma/\sigma,\\
L_1(0) &= L_2(0).
\end{aligned}
\tag{S2}
$$

In practice, we find these simplifying assumptions adequate for typical outbreak dynamics, but the initial conditions could also be included as independent parameters.

To connect the model equations to the data, we include a differential equation for the cumulative numbers of new infections $\bar{I}$ (with initial value 0).

$$d\bar{I}/dt = \quad 2\sigma L_2. \qquad (S3)$$

Then, from $\bar{I}(t)$, we calculate the numbers of actual infections $Y(t)$, reported cases $C(t)$, hospitalizations $H(t)$, and deaths $D(t)$ on day $t$, using the equation Eqs (S4).

$$\begin{aligned} Y(t) &= \kappa\big(\bar{I}(t) - \bar{I}(t-1)\big) \\ C(t) &= \kappa\varphi_{IC}\big(\bar{I}(t-\tau_{IC}) - \bar{I}(t-\tau_{IC}-1)\big), \\ H(t) &= \kappa\varphi_{IC}\varphi_{CH}\big(\bar{I}(t-\tau_{IC}-\tau_{CH}) - \bar{I}(t-\tau_{IC}-\tau_{CH}-1)\big), \\ D(t) &= \kappa\varphi_{IC}\varphi_{CD}\big(\bar{I}(t-\tau_{IC}-\tau_{CD}) - \bar{I}(t-\tau_{IC}-\tau_{CD}-1)\big). \end{aligned} \qquad (S4)$$

### Fitting models to data

We assume that daily reported cases, hospitalizations, and deaths are drawn from Poisson distributions with means $C(t)$, $H(t)$, and $D(t)$, respectively. The negative log likelihood $NLL_C(\theta)$ of observing the reported cases, $z_C(t)$ on each day $t$ in the set of days included for fitting cases, denoted $\{t_C\}$, is given in Eq (3). Here $\theta$ represents the estimated parameters for the Phase 2 or 3 models, as appropriate.

$$NLL_C(\theta) = \sum_{t\in\{t_C\}} C(t;\theta) + \sum_{t\in\{t_C\}} z_C(t)! + \sum_{t\in\{t_C\}} z_C(t) \times \log\big(C(t;\theta)\big) \tag{3}$$

The negative log-likelihoods for hospitalizations and deaths are defined analogously, using hospitalization and death data, $z_H(t)$ and $z_D(t)$, and their data periods $\{t_H\}$ and $\{t_D\}$, respectively. (The difference in data periods is discussed below). The total negative log-likelihood is the sum of the three negative log-likelihoods

$$NLL(\theta) = NLL_C(\theta) + NLL_H(\theta) + NLL_D(\theta) \tag{4}$$

This formulation gives weight to each data type commensurate with the number of observations, which will typically result in the optimizer prioritizing fits to the reported cases. As an advanced option, you may introduce weights on the three negative log-likelihoods to adjust how much priority is given to each data type.

Because of the burn-in period for case reporting by symptom onset date and the time between case reporting and hospitalizations and death, each of the three data streams needs to be informed by a different set of dates $\{t_C\}$, $\{t_H\}$, and $\{t_D\}$ corresponding, at least approximately, to the same population. For example, if there is a 2-week delay between case report and death on average, the trend in cases today will not show up in the death data for 2 weeks (Figure 1b). To account for the burn-in period $\omega$ and the delays $\tau_{CH}$ and $\tau_{CD}$, we define the data periods as follows as a function of today's date $t_0$ and the look-back period $\varepsilon$, i.e., the number of days we are looking back to inform the model fits.

$$\begin{aligned} \{t_C\} &= \{t_0 - \varepsilon, t_0 - \omega\} \\ \{t_H\} &= \{t_0 + \tau_{CH} - \varepsilon, t_0\} \\ \{t_D\} &= \{t_0 + \tau_{CD} - \varepsilon, t_0\} \end{aligned} \tag{7}$$

For our purposes, we assume that no burn-in is needed for hospitalizations and deaths, but a burn-in period can be added analogously for those data streams if needed. Because $\tau_{CH}$ and $\tau_{CD}$ change the optimization problem by changing $\{t_H\}$ and $\{t_D\}$, it is best to define them *a priori* (typically through a preliminary analysis of delays in trends) rather than optimize them along with other parameters. As $\tau_{IC}$ does not affect the optimization, it is not identifiable, and we recommend fixing it or sampling it from a reasonable range based on clinical information.

### "A large number" of parameter combinations

In each of the three Phases of the outbreak, our approach to uncertainty uses a sampling approach. Specifically, using Sobol' sampling (although other approaches, such as Latin hypercube sampling could also be used), we sample a large number of combinations of parameters from across their ranges, generate projections for each combination, and calculate credible intervals for each day in the projection. How many samples, exactly, is "a large number"? Unfortunately, there is no *a priori* number of combinations that will adequately cover the parameter space and achieve convergence in the quantities of interest. The answer depends on model complexity, the number of parameters to be sampled, and needed precision. In general, one starts with a smaller number and increases the sample size until the quantities of interest converge. Once a number is chosen, you don't necessarily need to repeat this exercise every time a new projection is generated, though it might be worthwhile to check if the answer changes when the dynamics qualitatively change.

Below, we include tables for each phase illustrating how some of the key quantities change as the number of parameter samples increases.

| Phase 1: Projections at six weeks | | | |
|---|---|---|---|
| Number of samples | 95% CI for cases | 95% CI for hospitalizations | 95% CI for deaths |
| 100 | (1,555; 15,221) | (20; 500) | (1; 30) |
| 500 | (1,520; 15,058) | (17; 541) | (1; 34) |
| 1,000 | (1,517; 15,104) | (17; 550) | (1; 33) |
| 5,000 | (1,516; 15,120) | (17; 551) | (1; 33) |
| 10,000 | (1,516; 15,126) | (17; 553) | (1; 33) |
| 50,000 | (1,516; 15,122) | (17; 556) | (1; 33) |
| 100,000 | (1,516; 15,122) | (17; 555) | (1; 33) |

| Phase 2: Projections at two weeks | | | |
|---|---|---|---|
| Number of samples | 95% CI for cases | 95% CI for hospitalizations | 95% CI for deaths |
| 100 | (4,122; 11,841) | (236; 729) | (37; 100) |
| 500 | (4,040; 12,060) | (235; 730) | (37; 100) |
| 1,000 | (4,084; 11,977) | (234; 736) | (37; 100) |
| 5,000 | (4,057; 11,999) | (233; 733) | (36; 101) |
| 10,000 | (4,048; 11,968) | (233; 735) | (36; 100) |
| 50,000 | (4,052; 11,978) | (233; 734) | (37; 101) |
| 100,000 | (4,053; 11,982) | (233; 733) | (37; 101) |

| Phase 3: Projections at six weeks | | | |
|---|---|---|---|
| Number of samples | Median (95% CI) for cases | Median (95% CI) for hospitalizations | Median (95% CI) for deaths |
| 100 | 11,166 (3,387; 29,036) | 851 (328; 1,680) | 113 (60; 164) |
| 500 | 11,170 (4,117; 30,338) | 852 (387; 1,740) | 113 (68; 168) |
| 1,000 | 11,404 (4,108; 30,413) | 864 (386; 1,744) | 114 (68; 168) |
| 5,000 | 11,747 (4,146; 30,247) | 883 (389; 1,735) | 115 (69; 168) |
| 10,000 | 11,973 (4,237; 30,425) | 894 (396; 1,738) | 116 (69; 168) |
| 50,000 | 11,923 (4,184; 30,370) | 893 (392; 1,738) | 116 (69; 168) |
| 100,000 | 11,942 (4,217; 30,370) | 894 (395; 1,738) | 116 (69; 168) |

For the purposes of our code, we have chosen 10,000 samples as adequate precision, especially up to the number of significant figures that would typically be reported.

One concern for Phase 3 is the comparatively slow speed of the two-step combination of sampling and estimation. On a typical machine, we were able to process about 5,600 samples per hour in serial. To improve computational speed, we provide (as the default option), code to run the process in parallel on a machine with multiple cores. With parallelization, processing the 10,000 samples takes approximately 10 minutes when using 10 cores. The specific timing will vary by the computational power of the machine.

### Threshold for “reasonable” fit to the data

In Phase 3, we want to ensure that all parameter sets included in the projections reasonably fit the data well. There may be combinations of the sampled parameters that cannot be made to fit the data well through tuning of the estimated parameters. How many parameter combinations need to be discarded will depend on the bounds of the sampled parameters. Modelers may want to investigate if certain values of sampled parameters consistently produce poor fits to the data; if so, tightening the interval of sampled parameters may improve computational efficiency.

Unfortunately, there is no *a priori* threshold that defines a “reasonable” fit. There are theoretical likelihood thresholds that one can use to develop confidence intervals for the best-fit trajectory, they are inappropriately narrow for our purpose of finding all trajectories that reasonably fit the recent data. Instead, discussion between the modelers and public health partners can build consensus about what threshold produces reasonable fits. We recommend framing this conversation without using technical language around likelihoods and thresholds but instead graphically compare the results generated by a few threshold options. Once the quality of “reasonable fit” has been agreed upon, the specific threshold can be tuned as needed.

Below, we illustrate the quality of fit to the data (and subsequent projections) for four thresholds: no threshold, 150% of the minimum NLL, 110% of the minimum NLL, and the 95th quantile of the chi-square distribution with 10 degrees of freedom, which produces the simultaneous confidence interval for the best-fit projection (Raue et al., 2009). Including all parameter combinations regardless of fit (no threshold), produces trajectories that do not represent the dynamics of the data. On the other side, the threshold producing the simultaneous 95% CI for the best-fit projection eliminates many trajectories that are plausible given the data to-date. Either of the other two examples could be appropriate. For the examples in this manuscript, we used 110% of the minimum NLL as our threshold.

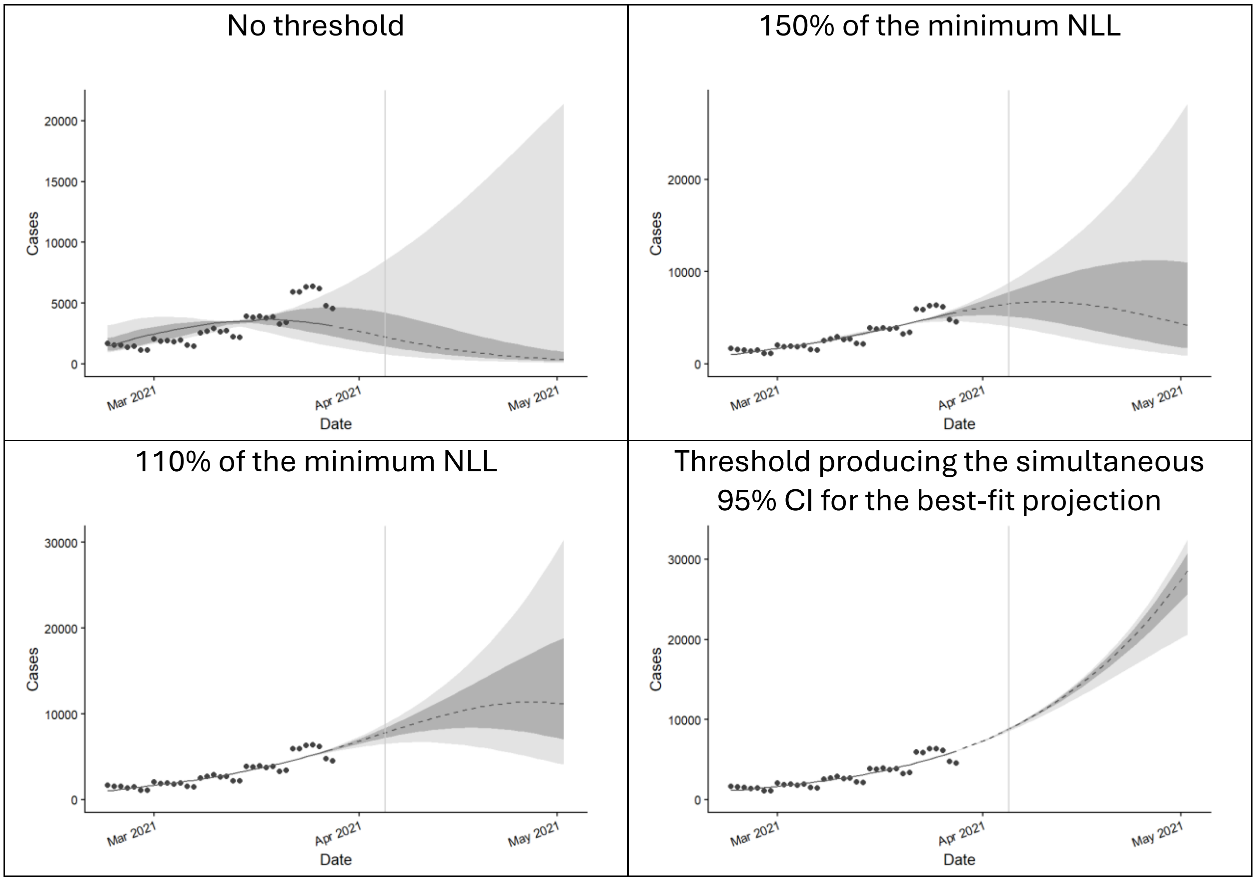
No threshold
150% of the minimum NLL
110% of the minimum NLL
Threshold producing the simultaneous 95% CI for the best-fit projection
Cases
Date
Mar 2021
Apr 2021
May 2021
0
5000
10000
15000
20000
30000
